## Supplementary Note 1&2, Supplementary Figures, Supplementary Table 1-2 for "The mediating role of mammographic density in the protective effect of early-life adiposity on breast cancer risk: a multivariable Mendelian randomization study"

In this document:

- Supplementary Tables S1 and S2
- Supplementary Note 1 (MR results using MD GWAS adjusted for BMI)
  - Supplementary Figures S1-S3
- Supplementary Figures S4-S9
- Supplementary Note 2 (Mediation analysis)

Supplementary Tables S3-S21, S22-S27, S28-S39 are provided in separate xlsx files.

Tables S3-S14 and S28-S29 are equivalent; the former contains results for the unadjusted MD data (as in the main results), while the latter results are based on MD data adjusted for BMI (as in [1]).

---

**Supplementary Table S1**. Summary of breast cancer GWAS datasets from the BCAC 2017 [2] and BCAC 2020 [3] releases. Sample sizes for BCAC 2020 were taken from [4].

| Release | Subtype | Receptor/grade status | Sample size | Cases | Controls | % cases |
|---|---|---|---|---|---|---|
| 2017 | Overall sample 2017 | | 228,951 | 122,977 | 105,974 | 53.7% |
| | ER+ | ER+ | 175,475 | 69,501 | 105,974 | 39.6% |
| | ER- | ER- | 127,442 | 21,468 | 105,974 | 16.9% |
| 2020 | Overall sample 2020 | | 247,173 | 133,384 | 113,789 | 54.0% |
| | Luminal A | ER+ and/or PR+, HER2−, grades 1 and 2; | 155,244 | 63,767 | 91,477 | 41.1% |
| | Luminal B1 | ER+ and/or PR+, HER2+; | 107,419 | 15,942 | 91,477 | 14.8% |
| | Luminal B2 | ER+ and/or PR+, HER2−, grade 3; | 107,419 | 15,942 | 91,477 | 14.8% |
| | HER2-enriched | ER− and PR−, HER2+ | 102,105 | 10,628 | 91,477 | 10.4% |
| | TNBC | ER− and PR−, HER2− | 100,079 | 8,602 | 91,477 | 8.6% |

**Supplementary Table S2.** The number of MD phenotypes' genetic instruments in unadjusted GWAS data (primary dataset) and adjusted for BMI data (dataset used for comparison only; corresponds to data published in [1]).

| | Unadjusted data | | | Adjusted for BMI data | | |
|---|---|---|---|---|---|---|
| | Total N after extraction | N in BCAC outcome 2017 / 2020 | | Total N after extraction | N in BCAC outcome 2017 / 2020 | |
| **Dense area** | 21 | 20 | 20 | 25 | 23 | 22 |
| **Non-dense area** | 8 | 7 | 8 | 16 | 15 | 16 |
| **Percent density** | 11 | 11 | 11 | 16 | 15 | 16 |

### Supplementary Note 1. MR results when using MD GWAS adjusted for BMI

1) Effect of childhood/adult body size on MD

When using the MD GWAS data adjusted for adult BMI (originally published in Sieh *et al* (2020) [1]), we observed an attenuation in the MVMR results (Supplementary Figure S1, Supplementary Table S30) of adult body size on all MD phenotypes, which may have suggested that childhood body size affects MD independently of adult body size.

This result, however, was a consequence of adjustment for BMI in the original GWAS summary statistics, leading to a double adjustment in MVMR with adult body size. This is a good example of how adjusted GWAS data in MR may create misleading results [5][6]. As such, we used the MD GWAS unadjusted for BMI for all primary analyses undertaken in this study.

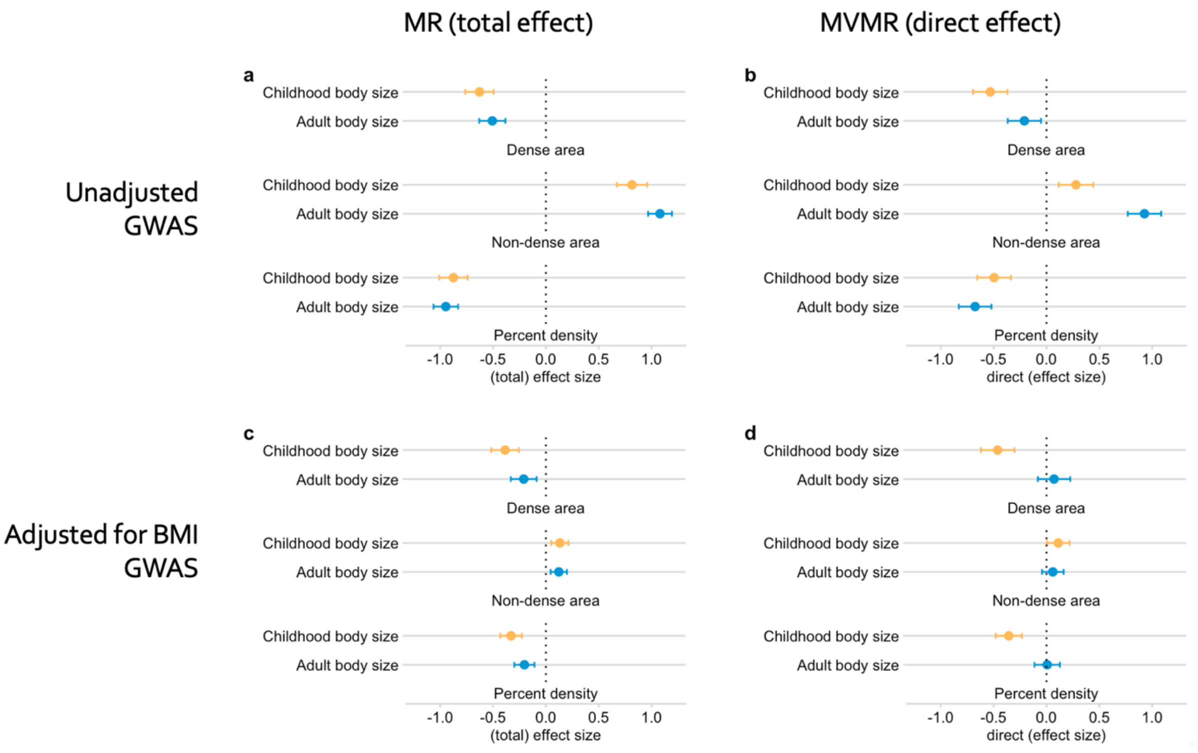


**Supplementary Figure S1.** The effect of childhood body size and adult body size on MD phenotypes (dense area, non-dense area, percent density), when using **unadjusted** MD GWAS (a, b) and **adjusted for BMI** MD GWAS (c, d). The results are presented for univariable MR total effect of each exposure on the outcome (a, c), and MVMR direct effect (b, d). Bars indicate 95% confidence intervals around the point estimates from IVW analyses.

2) Effect of age at menarche on MD

Age at menarche had no effect on MD (DA and PD) when using the MD GWAS adjusted for BMI (Supplementary Figure S2, Supplementary Table S32).

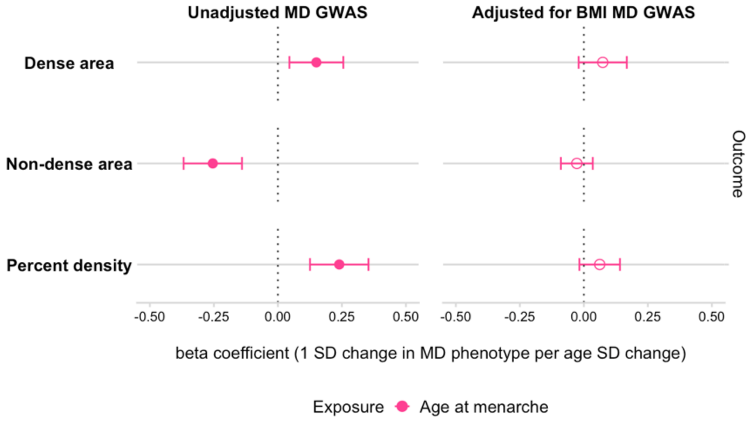


**Supplementary Figure S2**. The effect of age at menarche on MD phenotypes (dense area, non-dense area, percent density), when using **unadjusted** MD GWAS and **adjusted for BMI** MD GWAS. Bars indicate 95% confidence intervals around the point estimates from IVW analyses. The empty circle data points highlight the results where confidence intervals overlap the null.

### 3) Effect of MD on breast cancer

The comparison of results using the BMI-adjusted and unadjusted MD GWAS are presented in Supplementary Figure S3 and Supplementary Table S36. When using the adjusted data, a few more instruments were available for the analysis (Supplementary Table S2), however, there was similar heterogeneity among the instruments and imprecise estimates of effect on the majority of breast cancer subtypes.

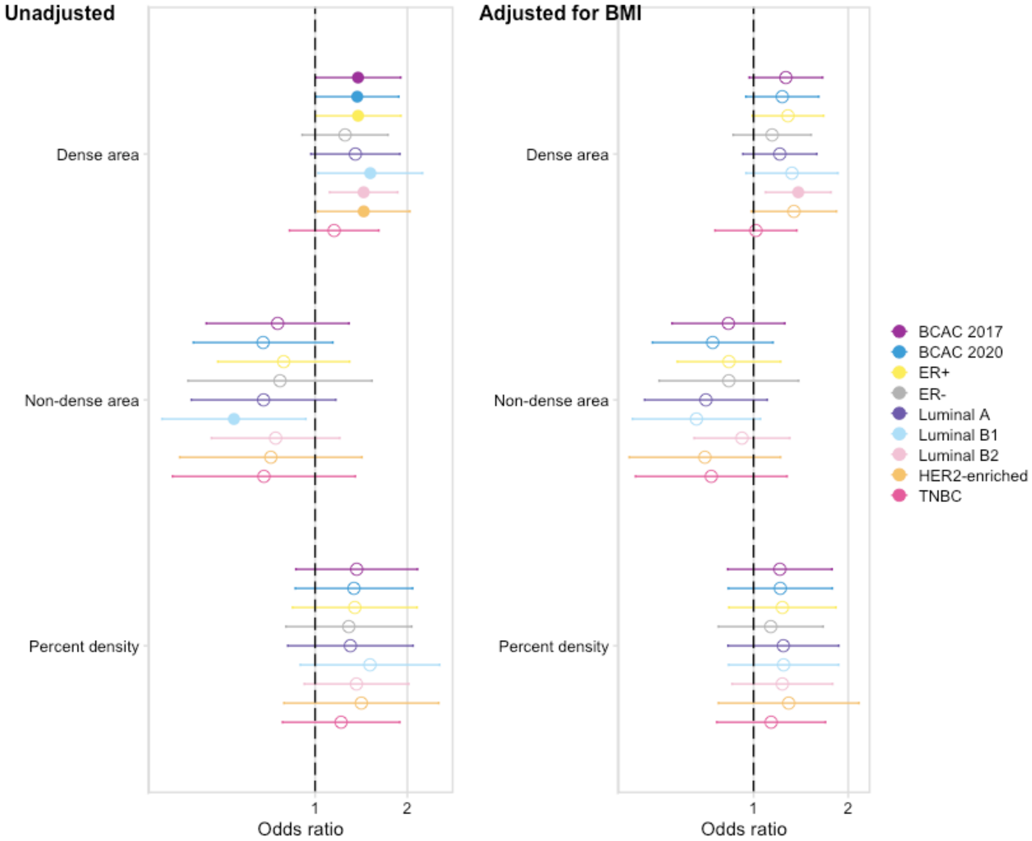


**Supplementary Figure S3**. The total (univariable MR) effect of MD phenotypes on breast cancer (overall and subtypes samples). **Unadjusted** plot (left) is the same as the results presented in Figure 3a in the paper, and is based on MD GWAS data unadjusted for BMI. **Adjusted for BMI** plot (right) is based on the data adjusted for BMI, as published in [1]. The instruments from both GWAS were extracted as described in Methods. The plots show the odds of breast cancer per SD higher MD phenotype. Bars indicate 95% confidence intervals around the point estimates from IVW analyses. The empty circle data points highlight the results where confidence intervals overlap the null.

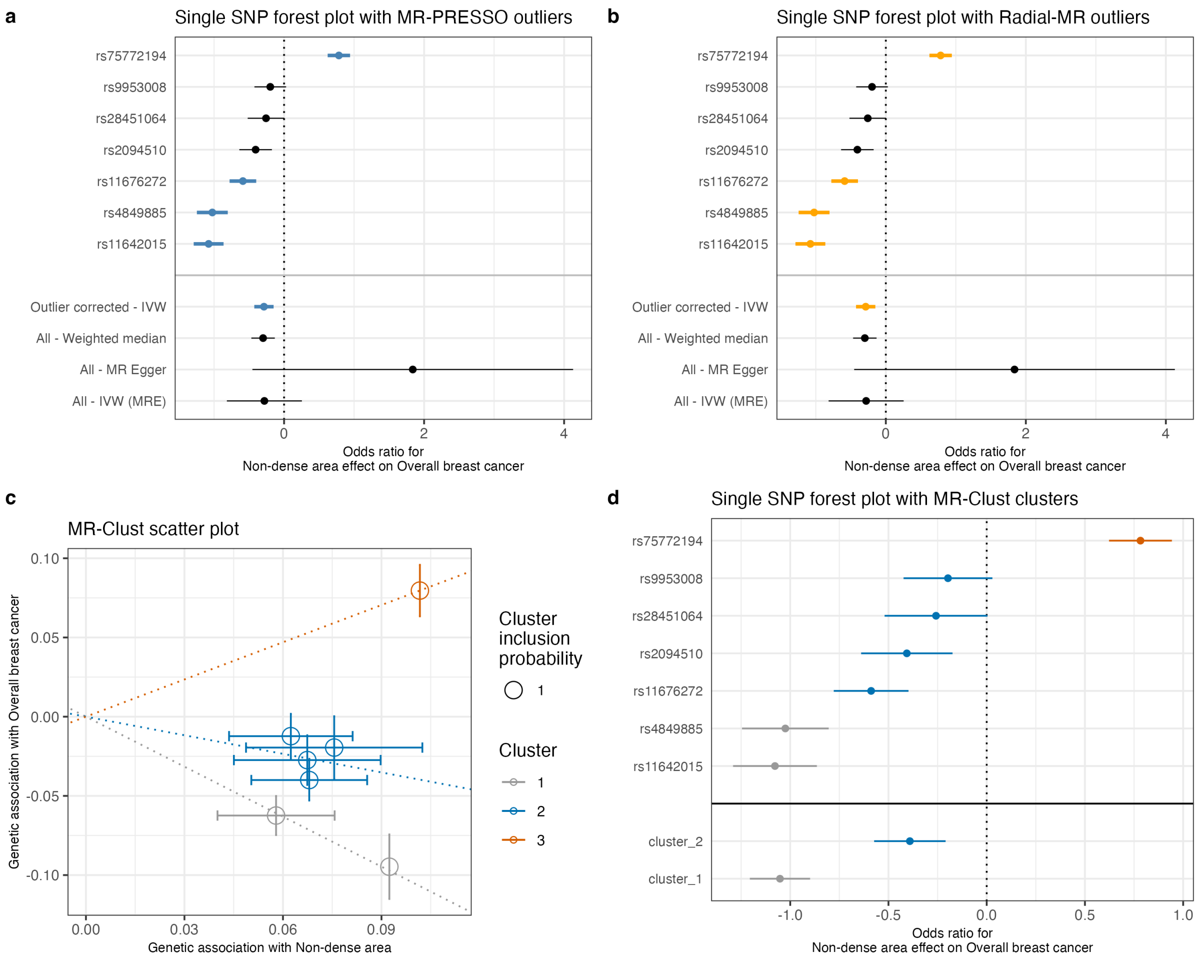


**Supplementary Figure S4. Exploring the heterogeneity of genetic instruments of non-dense area phenotype on breast cancer (BCAC 2017 overall sample). (a)** Single SNP forest plot (Wald Ratio estimates), with SNPs identified as outliers by MR-PRESSO marked in blue. The outlier corrected estimate is presented along with the standard MR methods estimates. **(b)** Single SNP forest plot with SNPs identified as outliers by Radial-MR marked in yellow. The outlier corrected estimate is presented along with the standard MR methods estimates. **(c)** MR-Clust scatter plot showing genetic association with dense area and breast cancer per SD change in dense area. Each genetic variant is represented by a point. Error bars are 95% confidence intervals of the Wald Ratio for each variant. Colours represent the clusters, and dotted lines represent the cluster means, the point size denotes cluster inclusion probability. The null cluster, coloured pink, relates to variants with null effect, whilst the black “junk cluster” are variants that were not assigned to any cluster. The error bars denote the standard error estimates of the Wald Ratio for each instrumental variable. **(d)** Single SNP forest plot with SNPs coloured by the cluster membership assigned by MR-Clust (using the same colours as in the scatter plot). The IVW MR estimates for each cluster are presented below single SNP estimates.

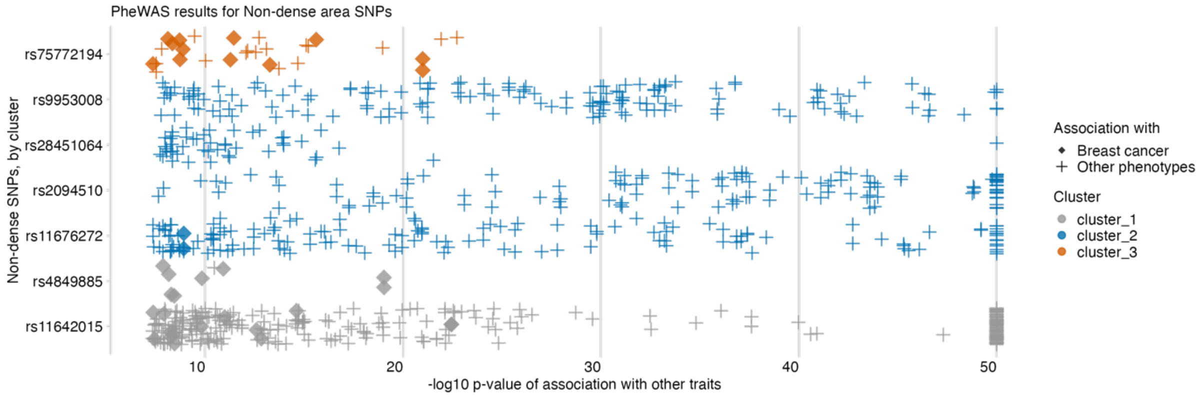


**Supplementary Figure S5. PheWAS results for non-dense area phenotype genetic variants, ordered by SNP effect and cluster membership** (from MR-Clust). The data points are other traits associated with a non-dense area SNPs (y-axis) at p-value <1e-08 (x-axis, -log10 scale, capped at value 50). The colour shows the cluster membership, in the same palette and order as in Figures S4c/S4d. Data points represented by solid ‘diamond’ shapes are breast cancer outcomes; ‘plus’ shapes are all other traits.

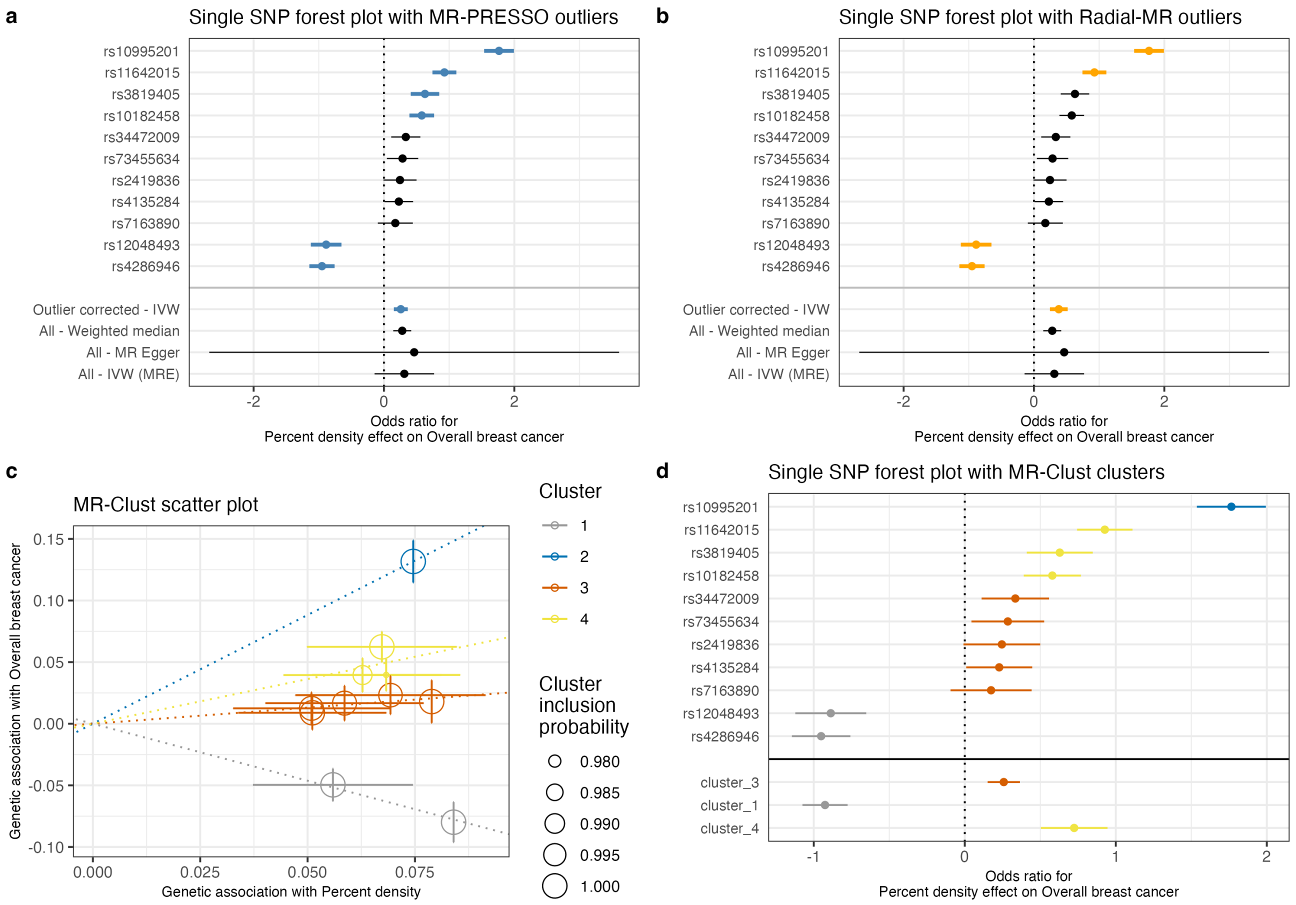


**Supplementary Figure S6. Exploring the heterogeneity of genetic instruments of percent density phenotype on breast cancer (BCAC 2017 overall sample). (a)** Single SNP forest plot (Wald Ratio estimates), with SNPs identified as outliers by MR-PRESSO marked in blue. The outlier corrected estimate is presented along with the standard MR methods estimates. **(b)** Single SNP forest plot with SNPs identified as outliers by Radial-MR marked in yellow. The outlier corrected estimate is presented along with the standard MR methods estimates. **(c)** MR-Clust scatter plot showing genetic association with dense area and breast cancer per SD change in dense area. Each genetic variant is represented by a point. Error bars are 95% confidence intervals of the Wald Ratio for each variant. Colours represent the clusters, and dotted lines represent the cluster means, the point size denotes cluster inclusion probability. The null cluster, coloured pink, relates to variants with null effect, whilst the black "junk cluster" are variants that were not assigned to any cluster. The error bars denote the standard error estimates of the Wald Ratio for each instrumental variable. **(d)** Single SNP forest plot with SNPs coloured by the cluster membership assigned by MR-Clust (using the same colours as in the scatter plot). The IVW MR estimates for each cluster are presented below single SNP estimates.

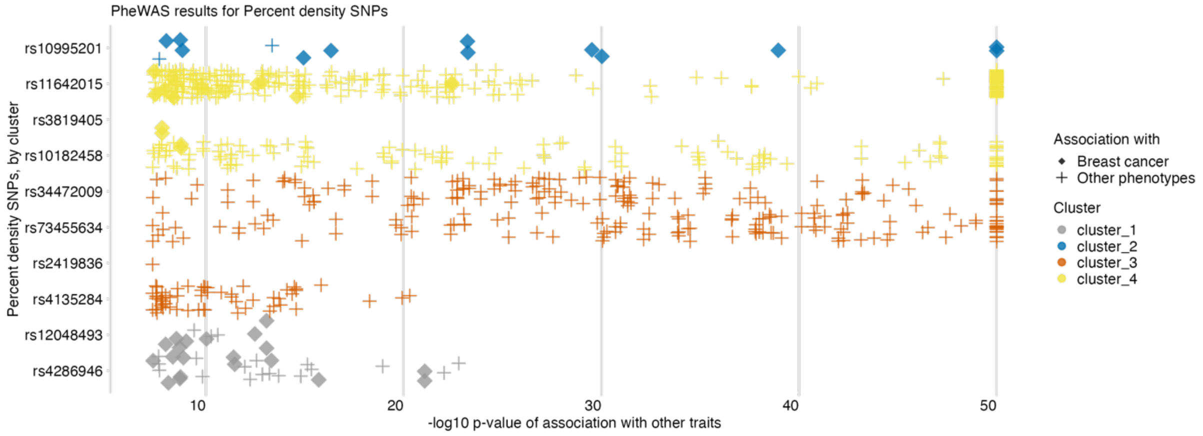


**Supplementary Figure S7. PheWAS results for percent density phenotype genetic variants, ordered by SNP effect and cluster membership** (from MR-Clust). The data points are other traits associated with a percent density SNPs (y-axis) at p-value <1e-08 (x-axis, -log10 scale, capped at value 50). The colour shows the cluster membership, in the same palette and order as in Figures S6c/S6d. Data points represented by solid 'diamond' shapes are breast cancer outcomes; 'plus' shapes are all other traits.

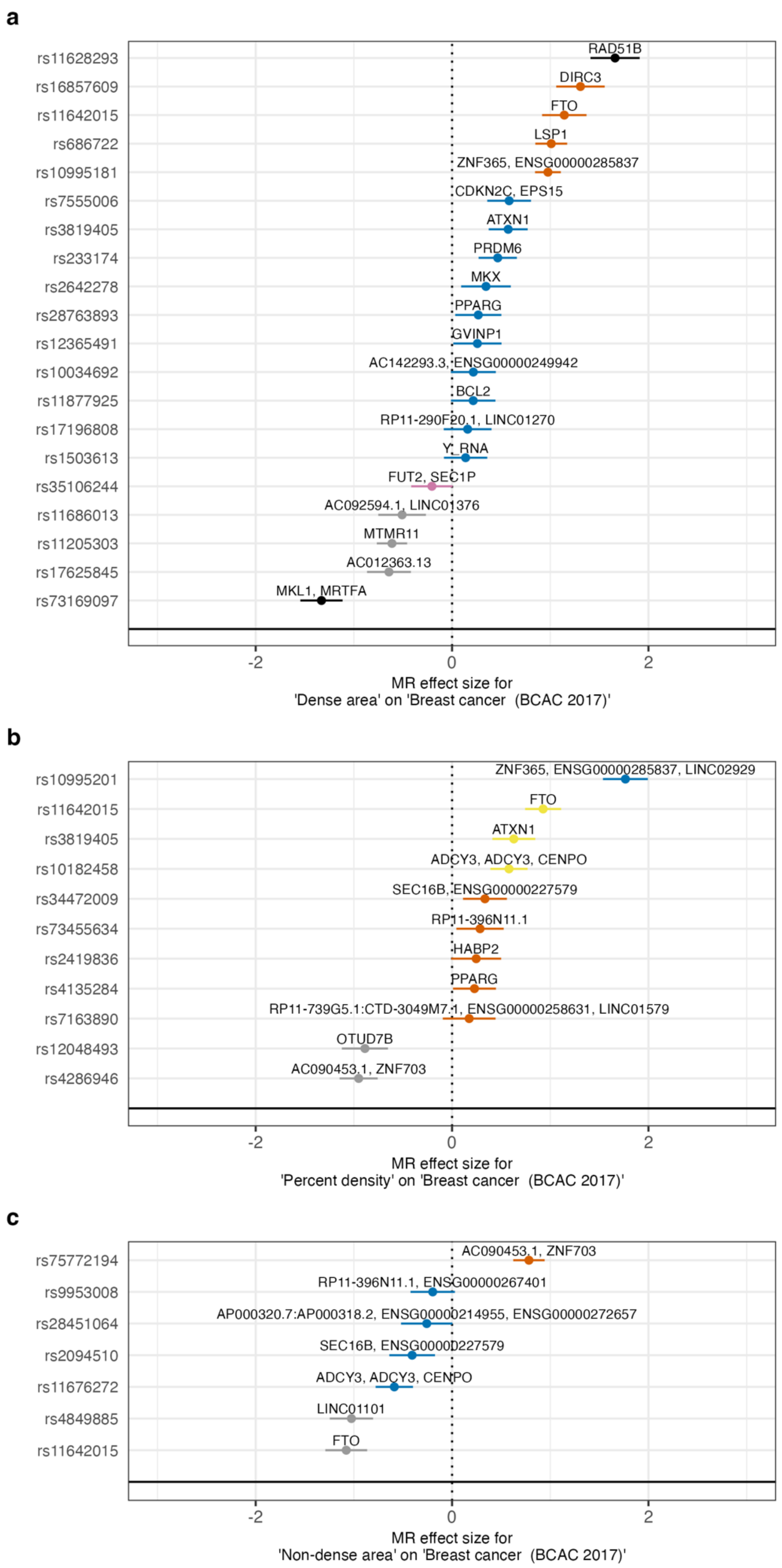


**Supplementary Figure S8.** (a) Dense area, (b) Percent density, (c) Non-dense area single SNP forest plot with SNPs coloured by the cluster membership assigned by MR-Clust, with each SNP mapped to a gene.

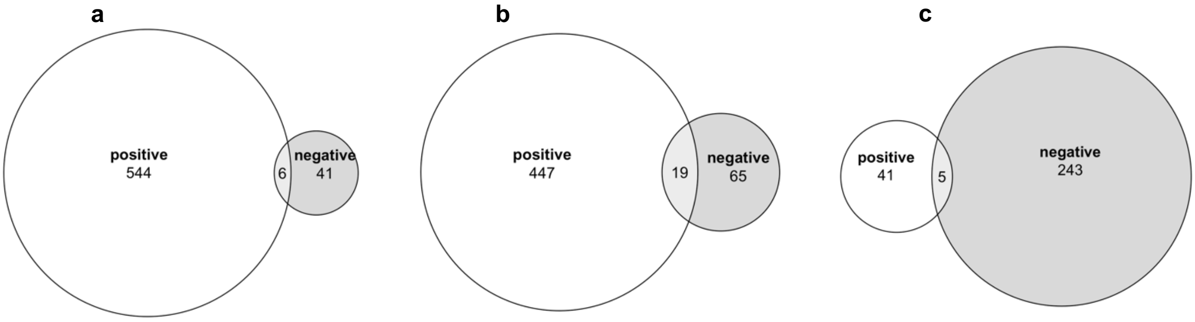


**Supplementary Figure S9.** (a) Dense area, (b) Percent density, (c) Non-dense area pathways counts Venn diagrams. For each phenotypes, instrument SNPs were mapped to genes and then pathways. SNPs/genes were splits into ‘positive’ and ‘negative’ effect direction sets, according to clustering results and individual SNPs estimates. The Venn diagrams show the number of unique pathways mapping to SNPs/genes in each effect direction set.

### Supplementary Note 2. Mediation analysis calculation

Mediation analysis was carried out for childhood body size (exposure), dense area (mediator), and breast cancer BCAC 2017 overall sample (outcome).

We estimated the proportion of childhood body size effect on breast cancer that is mediated via dense area.

### Analysis outline

See more details on mediation analysis in Supplementary Section E of our previous work (reference [7]).

1) Estimate 'indirect effect' of exposure on outcome via the mediator, using two methods:
   a. Difference method: **indirect effect = total effect – direct effect**
      ⇒ Estimate SE and CIs for the indirect effect using 'Propagation of Errors' method
      se.indirect = sqrt(se_total ^2 + se_direct^2)

   b. Product method: **indirect effect = exposure->mediator effect * mediator->outcome effect**
      ⇒ Estimate SE and CIs for the indirect effect using 'Delta' method
      se.indirect = sqrt(a_beta^2 * b_se^2 + b_beta^2 * a_se^2 ),
      where **a** is **exposure->mediator** and **b** is **mediator->outcome**

      **mediator->outcome effect** could be total effect or direct effect (from MVMR)

2) Estimate 'proportion mediated'
   ⇒ **proportion mediated = indirect effect / total effect *100%**

### Calculation

#### 1a. Difference method

Indirect effect: –0.411 – (–0.195) = **–0.215**

SE = sqrt(0.0702^2 + 0.117 ^2) = 0.136

lo_ci = –0.215 – 1.96 * 0.136= –0.48
up_ci = –0.215 + 1.96 * 0.136= 0.05

–0.22 [–0.48: 0:05]

#### 1b. Product method (using mediator->outcome direct effect)

Indirect effect: –0.62 * 0.37 = **–0.2294**

SE = sqrt(–0.62^2 * 0.069^2 + 0.37^2 * 0.068^2 ) = 0.0496

lo_ci = –0.2294 –1.96 * 0.0496 = –0.326
up_ci = –0.2294 + 1.96 * 0.0496= –0.132

–0.23 [–0.33: –0.13]

#### 2. Proportion mediated (% mediated)

*Product method*

% mediated = **–0.2294** / –0.411 * 100% = **55.8%**
% mediated (upper CI) = –0.326 / –0.411 * 100% = 79.3%
% mediated (lower CI) = –0.132 / –0.411 * 100% = 32.1%

**56% [32% - 79%]**

*Difference method*

% mediated = **–0.215** / –0.411 * 100% = **52.3%**

CIs for % mediated are not calculated here as the effect CIs overlap the null.
